# Incidence trends of nontuberculous mycobacterial pulmonary infections in Australia, Cambodia, Japan, Thailand, and the United States

**DOI:** 10.64898/2026.06.27.26356627

**Authors:** Atha R. Pradana, Melinda M. Ashcroft, Pichanee Watthanasiri, Rachel A. Mercaldo, Lisa Kawatsu, Eriko Morino, Socheatraksmey Ung, Christina Yek, Emilie Matsumoto-Takahashi, Felicia Goh, Kan Khemnak, Suphanat Wongsanuphat, Panithee Thammawijaya, Natthakan Tipkrua, Sujittra Pomchiangpin, Sokleaph Cheng, Kozo Morimoto, Surakameth Mahasirimongkol, D. Rebecca Prevots, Rachel M. Thomson

## Abstract

**BACKGROUND:** Nontuberculous mycobacteria (NTM) are environmental organisms increasingly causing chronic respiratory infection. Although NTM pulmonary infection is rising globally, most studies are single-country. This study evaluated temporal trends in pulmonary NTM incidence across Queensland (Australia), Phnom Penh (Cambodia), Japan, Thailand, and the United States (US), and described regional species distribution.

**METHODS:** Laboratory and insurance claims data were used. Incident infections were defined using region-specific criteria. For Queensland, Japan, and Thailand, data and denominators covered entire regions. US estimates included Medicare beneficiaries aged ≥65 years, and Cambodian incidence was estimated from Phnom Penh data and standardised nationally. Incidence rates per 100,000 population and incidence rate ratios (IRRs) were calculated overall and by sex. Age-stratified analyses and species distributions were summarised where data were available.

**RESULTS:** Pulmonary NTM incidence increased in all regions and was highest in Japan (47.20–57.40 per 100,000) and lowest in Phnom Penh (0.23–0.38). Queensland showed the largest increase over 24 years (IRR 7.06, p<0.0001). Female predominance occurred in high-income regions, whereas Thailand showed ~1.5-fold male predominance and Phnom Penh showed no sex predominance. Incidence was higher among individuals aged ≥60 years. *Mycobacterium avium* complex predominated in Japan and Queensland, and *M. abscessus* in Thailand and Phnom Penh.

**CONCLUSIONS:** Pulmonary NTM incidence increased in all regions, varying by demographic patterns and species distribution. Differences largely reflect under-ascertainment related to diagnostic capacity and tuberculosis-focused health systems rather than true infection burden. Strengthened surveillance and diagnostic capacity are needed to define the global burden of NTM pulmonary infection.

**KEY MESSAGES:**

- Pulmonary NTM incidence increased across all five study regions, however varied substantially by region, age, sex, and species.
- This study provides the first NTM incidence estimates from Thailand and Cambodia, where limited diagnostic capacity and tuberculosis-focused health systems influence observed incidence.
- Combining laboratory and claims data across high- and middle-income regions provides robust, comparable estimates to inform NTM surveillance and public health planning.

## INTRODUCTION

Nontuberculous mycobacteria (NTM) are environmental mycobacteria with >200 species identified.^1^ NTM represent all mycobacteria except the *Mycobacterium tuberculosis* complex (MTBC), *M. leprae*, and *M. lepromatosis*.^2,3^ Most NTM infections are pulmonary (70–80%), with the remainder extrapulmonary or disseminated disease.^4,5^ These opportunistic pathogens are ubiquitous in the environment,^6^ inhabiting soils, sediments, dust, and natural waters.^7^ Transmission occurs through ingestion, inhalation, direct inoculation, and dermal contact,^2,8^ and may also occur via fomites.^9^

A global systematic review found that 82.1% of included studies reported increasing rates of NTM pulmonary infection, while 66.7% reported increasing rates of pulmonary disease (NTM-PD).^10^ Infection and disease are distinct: infection is defined through positive culture, whereas disease classification requires additional clinical, radiographic, and microbiologic evidence.^8,9^ In many high-income regions, incidence of NTM pulmonary infection and NTM-PD has increased steadily, including in Queensland (Australia)^11^, the United States (US)^12^, and Japan^13^. In contrast, population-based studies remain extremely limited in low- and middle-income countries (LMICs).^14,15^

The global literature on NTM epidemiology remains fragmented, with most studies focusing on single regions. Surveillance is limited because NTM infection is not notifiable in most countries, and awareness among clinicians and microbiologists remains low.^10^ In many LMICs, particularly those with endemic tuberculosis (TB), health systems remain heavily oriented toward TB control, which may overshadow recognition of NTM infection.^16^ Diagnostic capacity for mycobacterial culture and speciation is also constrained.^16^ International comparisons are further complicated by differences in surveillance systems, diagnostic approaches, and case definitions.^14^ Population age and sex distributions also vary across countries and may influence observed incidence. Comparative descriptive epidemiology across multiple regions is therefore needed to better characterise global pulmonary NTM incidence.

In this study, we analysed NTM data from five regions: Queensland (Australia), Phnom Penh (Cambodia), Japan, Thailand, and the US. We aimed to evaluate temporal trends in pulmonary NTM incidence, including sex- and age-specific patterns, and to describe variation in species distribution across regions.

## METHODS

### Study populations

#### Australia

NTM infections in Queensland, Australia are notified to the Queensland Health Notifiable Conditions (NoCS) Database.^11,17^ All NTM notifications in Queensland from 2001–2024 were included. An incident case was defined as the first positive NTM culture for an individual. A new incident case was recorded if a subsequent positive culture occurred more than 12 months after the initial specimen collection, regardless of the number of positive cultures during that period. For individuals with multi-species infection, each species was counted as an incident case. De-identified patient demographic data included age at specimen collection, sex, residential location and Indigenous status. Where available, site of infection and specimen type were used to classify infections as pulmonary or extrapulmonary. Incidence rates were calculated per 100,000 Queensland population using Australian Bureau of Statistics mid-year estimated resident population data, stratified by age and sex.^18^ Speciation methods changed during the study period. Matrix-Assisted Laser Desorption/Ionization Time-of-Flight (MALDI-TOF) was introduced in 2019, and in 2021 PCR genotyping databases were updated, and *M. fortuitum* was reported as *M. fortuitum* group (MFG). For consistency, all *M. fortuitum, M. peregrinum, M. fortuitum* complex species were grouped as MFG. From 2021, most notifications of *M. avium* complex (MAC) reflected identification at the complex level, or MAC species other than *M. avium* or *M. intracellulare*.^19^

#### Cambodia

Positive NTM data from 2010 to 2023 were extracted from the Laboratory Information System (LIS) of the Mycobacteriology Unit, Medical Biology Laboratory, Institut Pasteur du Cambodge, the only laboratory in Cambodia performing NTM culture and speciation. Case identification occurred through self-referral or hospital referral for suspected mycobacterial infection, including TB. Testing was conducted at a single centre in Phnom Penh and is therefore not nationally representative. Speciation methods evolved during the study period. From 2020, NTM speciation was performed for all culture-positive isolates using Bruker MALDI-TOF Biotyper. Prior to 2020, speciation was not consistently performed. This dataset included de-identified patient demographic information (age, sex, and residential province) and NTM species identification. An incident case was defined as an individual with one or more positive NTM cultures, with only one isolate included per calendar year. Incidence rates were calculated per 100,000 Cambodian population estimates from the United Nations World Population Prospects 2024.^20^ Incidence was stratified by sex; age-specific denominators were unavailable for further stratification by age.

#### Japan

NTM data were obtained from two sources, insurance claims data and laboratory-based surveillance data. Insurance claims NTM data from the National Health Insurance Claim Database (NDB) were analysed for 2010 to 2019. An incident case was defined as an International Classification of Diseases 10th Revision (ICD-10) code A31.0 (pulmonary mycobacterial infection, excluding leprosy and tuberculosis) or A31.9 (mycobacterial infection, unspecified), provided there had been no claim for either code in the preceding two years. Incidence rates were calculated using annual population estimates (October of each year, 2012-2019) from the Statistics Bureau of Japan and were stratified by age and sex.^21^ Laboratory-based surveillance defined incident NTM pulmonary infection as a single positive culture result from sputum, bronchial washing or lavage, transbronchial biopsy or lung biopsy specimens. Incidence date was defined as the first registration of NTM infection regardless of the number of NTM species detected.^13^ Incidence rates were estimated using the ratio of NTM pulmonary infection cases to newly notified pulmonary TB cases, a method previously established in Japan to estimate pulmonary NTM incidence. These approaches differ in methodology, with claims data providing population-based estimates and laboratory-based estimates derived using extrapolation, and are not directly comparable.

#### Thailand

NTM infection data were collected from electronic medical records submitted to the national Health Data Center (HDC) under the Ministry of Public Health (MOPH), which compiles data from healthcare facilities encompassing approximately 71.2% of the Thai population in 2024.^22^ Pulmonary NTM infections were identified using physician diagnoses coded as ICD-10 A31.0 recorded from 2014 through 2024. Only the initial diagnosis was included, follow-up visits within one year were excluded. Laboratory data from 2014 to 2024 were obtained from the Office of Disease Prevention and Control Region 5 (ODPC 5), which covers eight provinces in western Thailand: Kanchanaburi, Suphan Buri, Nakhon Pathom, Ratchaburi, Samut Sakhon, Samut Songkhram, Phetchaburi, and Prachuap Khiri Khan. As part of routine care, specimens from patients with positive acid-fast bacilli (AFB) staining results were analysed using molecular methods targeting both MTBC and NTM. Species identification of NTM was also performed using a line probe assay upon clinician request; consequently, some NTM-positive cases lacked species-level identification. Extracted variables included demographic characteristics (sex and age at diagnosis), clinical information (type of specimen, AFB staining result, and site of infection), and NTM species identification. Incidence rates per 100,000 population were calculated using annual population estimates by age and sex from the Civil Registration Statistics of the Department of Provincial Administration, Ministry of Interior.^23^

#### United States

NTM case data were obtained from the Medicare Carrier B claims dataset provided by the Centers for Medicare and Medicaid Services (CMS)^24^. Medicare beneficiaries are nationally representative of persons ≥65 years in the US.^25^ Incident NTM pulmonary infection cases were defined as claims with either ICD-9 code 031.0 (pulmonary diseases due to other mycobacteria) or ICD-10 code A31.0 recorded between 2010 and 2019, following a 24-month period without NTM claims. In this study, a single-claim definition was used to maximise case capture. Incidence rates were calculated per 100,000 population, using the total beneficiary population as the denominator.^12^

Across datasets, incidence reflects the underlying data source, with laboratory-based systems capturing microbiologically defined NTM infection and administrative claims data capturing clinically recognised NTM-PD.

### Statistical analysis

Incidence rates were calculated per 100,000 study population. Temporal trends were assessed using incidence rate ratios (IRRs) comparing the first and final years of the study period for each region. IRRs were estimated using unconditional maximum likelihood with Wald confidence intervals (*epitools* v0.5.10.132). Analyses were conducted in R (v4.5.0).

## RESULTS

### Trends in NTM pulmonary infection incidence

Across all five regions, pulmonary NTM incidence increased over time (Table 2; Figure 1). Incidence estimates differed by data source, with administrative claims data (ICD-coded claims: Japan [NDB], Thailand, and the US [CMS]) generally yielding lower estimates than laboratory data (Queensland [Australia], Phnom Penh [Cambodia], and Japan), reflecting differences in case definitions and ascertainment.

**Table 1.** Summary of NTM epidemiology methods and study populations by region.

| Geographic region | Data sources | Case definition | Study Period | Notes |
| --- | --- | --- | --- | --- |
| QLD, Australia | - QLD Health NOCS<br>- QMRL | - Culture-positive NTM isolate<br>- One isolate per person per year<br>- Multi-species infections counted separately | 2001-2024 | - Population-level laboratory surveillance<br>- Species identification via QMRL<br>- Includes species typically considered non-pathogenic |
| PNH, Cambodia | - Persons screened for suspected TB or NTM.<br>- Medical Biology Laboratory, Institut Pasteur du Cambodge. | - $\geq 1$ positive NTM culture<br>- One isolate per person per calendar year | 2010-2023 | - Single testing centre<br>- Incidence standardised to national population<br>- Species identification available from 2020-2023 |
| Japan | National Health Insurance Claim Database (NDB) | Pulmonary NTM infection identified using ICD codes A31.0 or A31.9 | 2012-2019 | - Administrative claims dataset capturing clinically recognised NTM infection |
|  | Private laboratories in Tokyo, Japan (SRL, Inc. and LSI Medience Corporation) | Positive NTM culture from: 1) a single positive culture from sputum, or 2) a positive culture from bronchial washing or lung biopsy specimen | 2015-2019 | - NTM species data from 2015<br>- Sex and age data were missing for 10-15% of cases; those records were excluded from sex- and age-specific incidence estimates, so numerators may not sum to the total |
| Thailand | National Electronic Medical Records (MOPH (HDC)) | $\geq 1$ ICD-10 code A31.0 recorded within a year | 2014-2024 | - Administrative claims covering approx. 71% of the Thai population<br>- Species-level data unavailable |
|  | Microbiology: speciation (ODPC5) | NTM identified from specimens of suspected TB patients | 2013-2024 | - Regional laboratory dataset<br>- Species-level identification was available for a subset of isolates |
| United States | Medicare Carrier B claims dataset (CMS) | - One claim of ICD 031.0 or A031.0<br>- No claims in the preceding two years | 2010-2019 | - Medicare beneficiaries aged $\geq 65$ years<br>- Species data unavailable |

**Abbreviations:** CMS, Centers for Medicare and Medicaid Services; HDC, Health Data Center; ICD, International Classification of Diseases; IOM, International Organization for Migration (Cambodia); MOPH, Ministry of Public Health (Thailand); NDB, National Health Insurance Claims Database; NOCS, Notifiable Conditions System; NTIP, National Tuberculosis Information Program; NTM, Nontuberculous mycobacteria; ODPC5, Office of Disease Prevention and Control Region 5; PNH, Phnom Penh; QLD, Queensland; QMRL, Queensland Mycobacterial Reference Laboratory; TB, tuberculosis.

**Table 2.** Incidence of NTM pulmonary infection by regions and study population.

| Geographic Region | IR<sup>#</sup> | Average annual IR<sup>#</sup> | AAPC | Date Range | IRR | 95% CI | *p*-value |
| --- | --- | --- | --- | --- | --- | --- | --- |
| QLD, Australia | 4.37-30.87 | 20.22 | 6.36 | 2001-2024 | 7.06 | 6.39-7.83 | < 0.0001 |
| PNH, Cambodia | 0.23-0.38 | 0.29 | 1.05 | 2010-2023 | 1.65 | 0.96-2.90 | 0.0548 |
| Japan (NDB data) | 39.40-43.80 | 42.20 | 1.33 | 2012-2019 | 1.11 | 1.06-1.16 | < 0.0001 |
| Japan (Lab data) | 47.20-57.40 | 53.12 | 4.75 | 2015-2019 | 1.22 | 1.17-1.26 | < 0.0001 |
| Thailand | 0.69-1.61 | 1.12 | 6.91 | 2014-2024 | 2.33 | 1.75-3.14 | < 0.0001 |
| US (CMS data) | 23.77-34.13 | 29.67 | 4.45 | 2010-2019 | 1.44 | 1.36-1.51 | < 0.0001 |

<sup>#</sup>Per 100,000 population. Abbreviations: AAPC, average annual percent change; CI, confidence interval; CMS, Centers for Medicare and Medicaid Services; IR, incidence rate; IRR, incidence rate ratio; NDB, National Health Insurance Claims Database; NTM, nontuberculous mycobacteria; PNH, Phnom Penh; QLD, Queensland.

**Figure 1.**
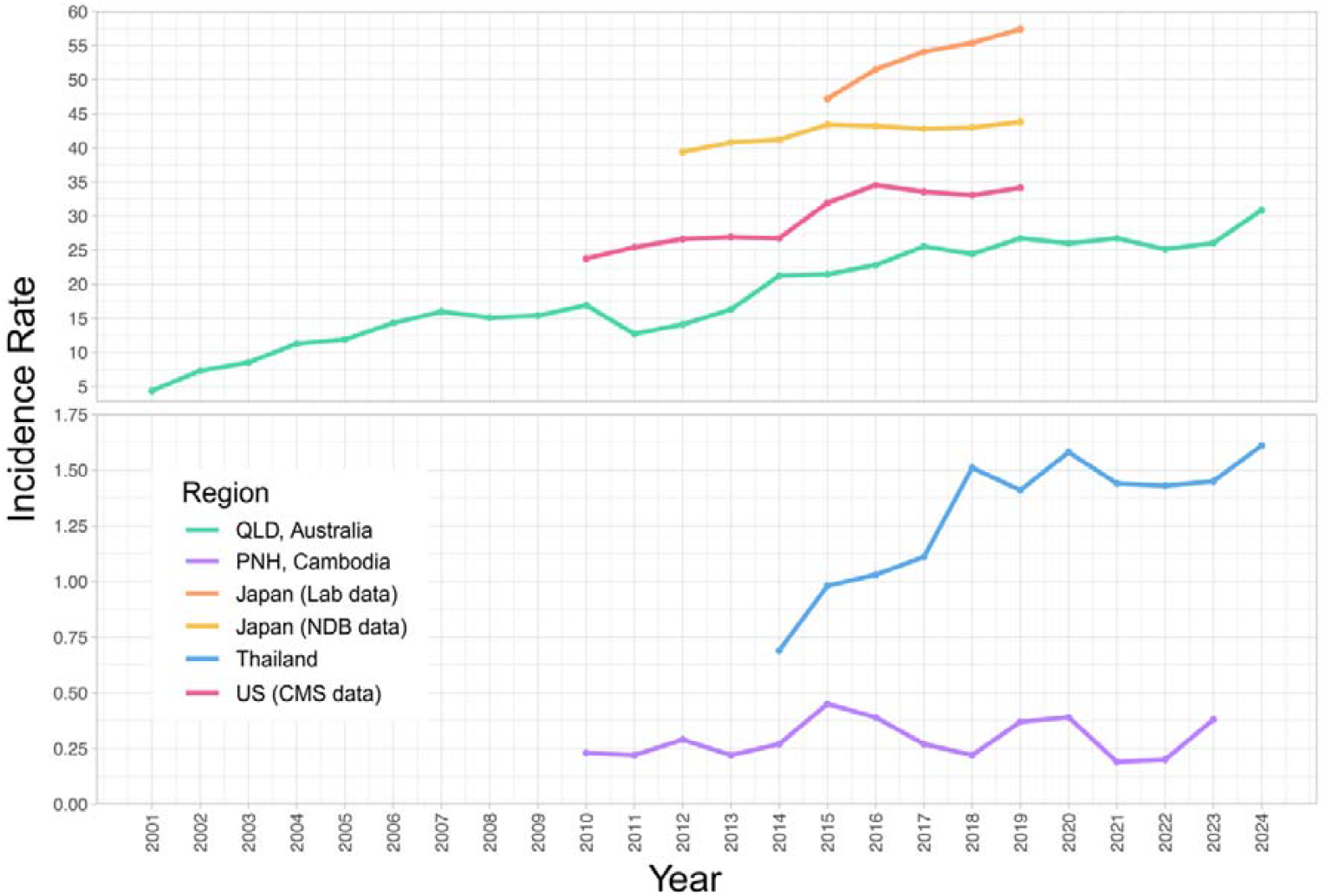
NTM pulmonary infection incidence per 100,000 population. Lines are coloured by region (see legend). Abbreviations: CMS, Centers for Medicare and Medicaid Services; NDB, National Health Insurance Claim Database; PNH, Phnom Penh; QLD, Queensland.

#### Administrative claims data

Incidence derived from NDB data in Japan was highest overall, ranging from 39.40–43.80 per 100,000 population and showed a modest but steady increase (IRR: 1.11; 1.33% average annual increase). In the US, incidence ranged from 23.77–34.13 per 100,000 and increased steadily (IRR: 1.44; 4.45% average annual increase). In Thailand, incidence was substantially lower (0.69–1.61 per 100,000), but showed the largest proportional increase (IRR: 2.33; 6.91% average annual increase).

#### Laboratory data

Laboratory-based incidence in Japan was highest overall (47.20–57.40 per 100,000), increased steadily (IRR: 1.22; 5.31% average annual increase), and consistently exceeded claims-based estimates. In Queensland, Australia, incidence varied substantially across study years (4.37–30.87 per 100,000 population) and increased markedly over time (IRR: 7.06; 6.36% average annual increase). In Phnom Penh, Cambodia, where incidence was standardised to the national Cambodian population, incidence was very low (0.23– 0.38 per 100,000 population) and fluctuated over time (IRR: 1.65; 1.05% average annual increase).

### Sex-specific trends in pulmonary NTM incidence

Sex-specific patterns of pulmonary NTM incidence differed by region and data source (Figure 2), with female predominance in high-income regions and a male predominance in Thailand.

**Figure 2.**
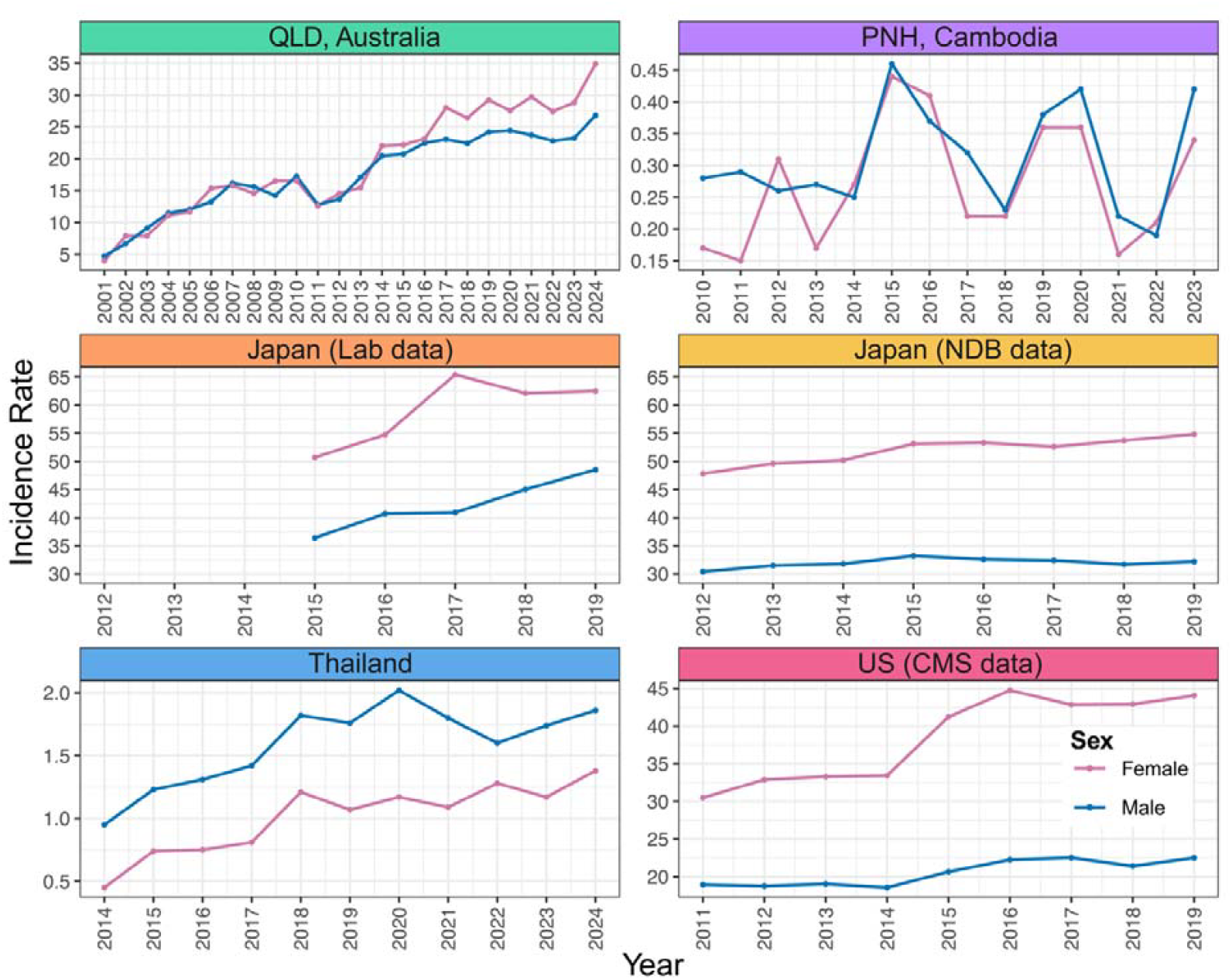
Pulmonary NTM incidence rates (IR) per 100,000 population by sex and region. Lines are coloured by sex (red, females; blue, males). Abbreviations: CMS, Centers for Medicare and Medicaid Services; NDB, National Health Insurance Claims Database; PNH, Phnom Penh; QLD, Queensland.

#### Administrative claims data

In Japan (NDB data) and the US (CMS data), females had higher incidence than males. In Japan, incidence ranged from 47.80–54.80 per 100,000 population in females and from 30.40–33.20 in males. In the US, incidence ranged from 30.48–44.75 per 100,000 population in females and from 18.94–22.51 in males. In contrast, Thailand showed consistently higher incidence in males throughout the study period, ranging from 0.95–2.02 per 100,000 population compared with 0.45–1.38 in females, corresponding to an approximately 1.5-fold difference.

#### Laboratory data

In Japan, laboratory-based estimates showed a similar sex-specific pattern to that observed in the NDB, with consistently higher incidence in females (50.70–65.40 per 100,000 population) than males (36.40– 48.50). In Queensland, Australia, incidence for males and females was comparable between 2001 and 2013 (females 4.01–16.55 per 100,000 vs. males 4.72–17.28), before shifting toward a clear female predominance from 2014 onwards. In Phnom Penh, Cambodia, incidence increased over time for both sexes (females 0.15–0.44 per 100,000 population vs. males 0.19–0.46), with marked year-to-year variability. In 2010, incidence in males was nearly double that of females, but by 2014 this difference had disappeared, with no clear sex predominance after.

### Age-stratified sex-specific trends in pulmonary NTM incidence

Age-stratified patterns differed across regions, with consistently higher incidence in individuals aged ≥60 years (Figure 3). Comparisons also varied by data source, with distinct patterns across administrative claims and laboratory datasets. Cambodia was not included because age-specific denominators were unavailable.

**Figure 3.**
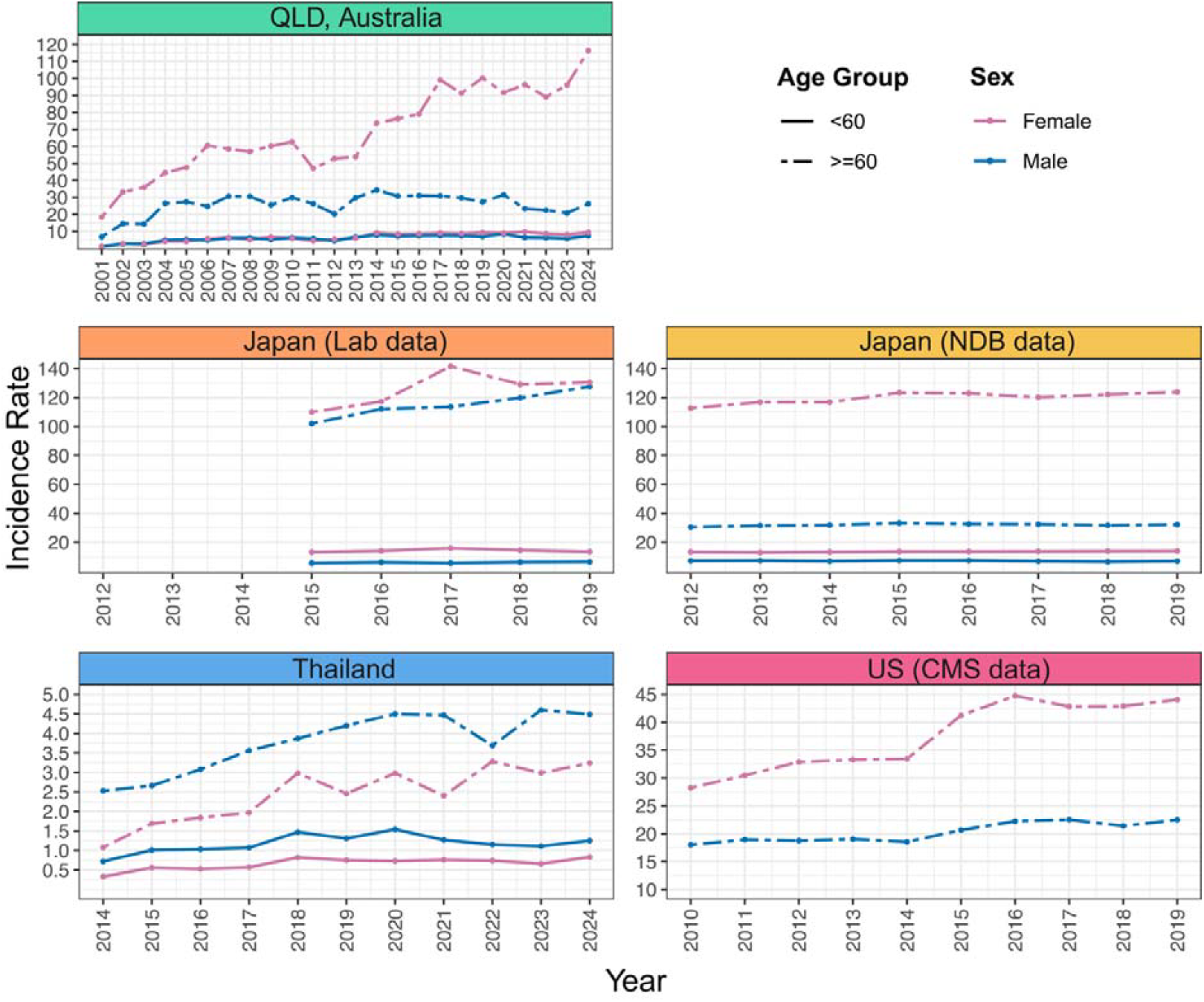
Pulmonary NTM incidence by sex, age group, and region. Solid lines represent those aged <60, while dashed lines represent those aged ≥60. Lines are coloured by sex (pink for females; blue for males). Cambodia was excluded due to unavailable age-specific denominators. Abbreviations: CMS, Centers for Medicare and Medicaid Services; NDB, National Health Insurance Claim Database; QLD, Queensland.

**Figure 4.**
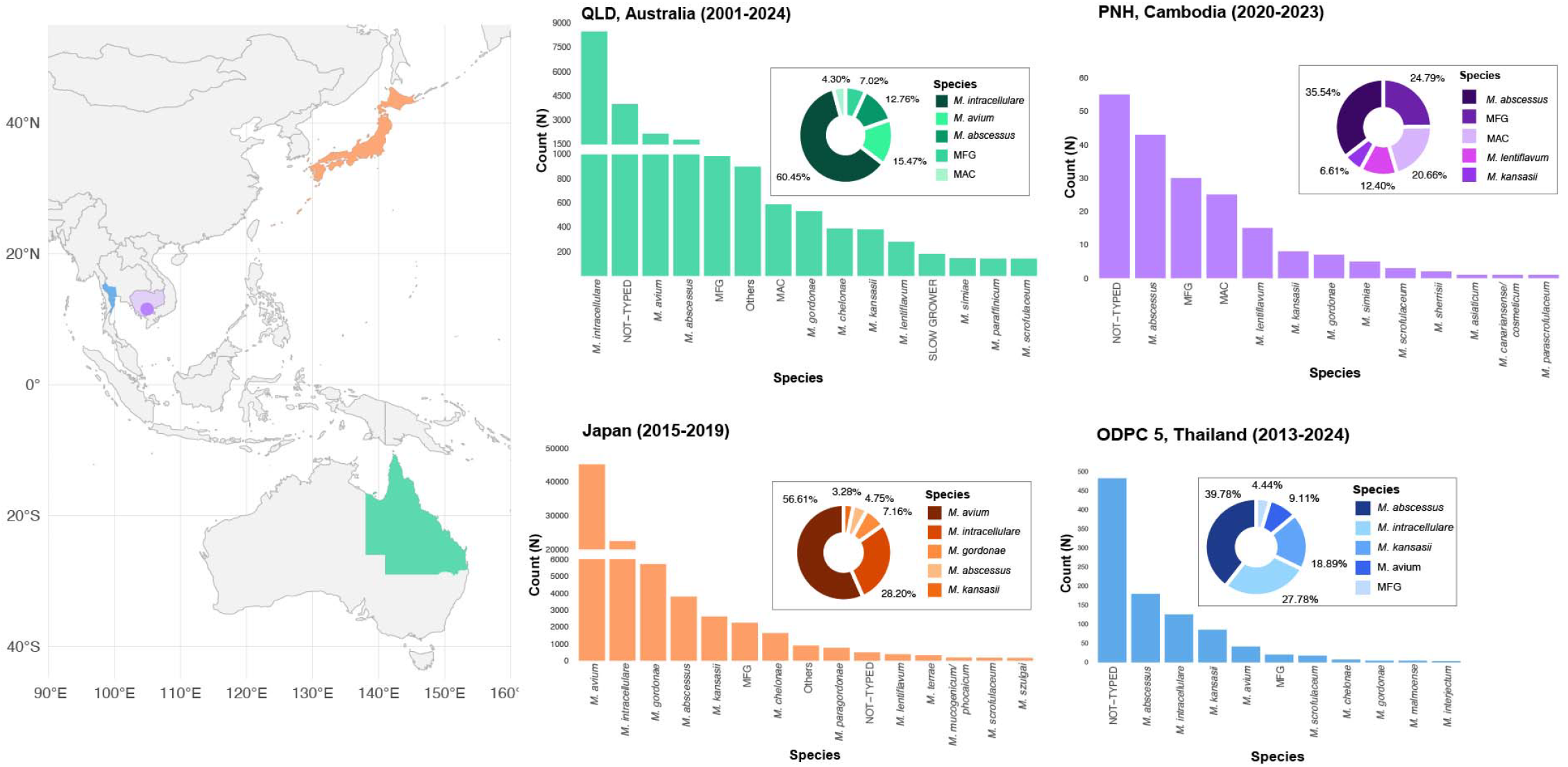
Distribution of nontuberculous mycobacterial (NTM) species by region. Bar graphs show the absolute number of isolates by species, and pie charts show the relative proportions of the top five species. All identified species, including isolates without species-level identification (“NOT-TYPED”), are included. For Phnom Penh, Cambodia and the ODPC 5 region of Thailand, all detected species are shown. For Queensland (QLD), Australia and Japan, the 14 most frequently isolated species are shown, with remaining species grouped as “Others”. Percentages in the pie charts are calculated using the total number of isolates among the top five species. The United States was excluded due to unavailable species-level data. In Queensland, MAC includes *M. chimaera* and other MAC species. In Japan, *M. intracellulare* may also include *M. chimaera*, depending on laboratory reporting practices. Abbreviations: MAC, *M. avium* complex; MFG, *M. fortuitum* group.

#### Individuals aged <60 years

Among individuals aged <60 years, trends varied across regions. In Japan, NDB-derived incidence closely paralleled laboratory estimates in females. Laboratory-based rates exhibited modest variability (13.0–15.80 per 100,000 population), whereas NDB estimates remained stable (13.10–13.80 per 100,000). A similar pattern was observed in males, where NDB-derived incidence remained stable (6.40–7.30 per 100,000) and laboratory-based estimates followed a slightly lower but comparable trajectory (5.50–6.30 per 100,000). In Queensland, Australia, incidence among females and males (1.14–9.69 per 100,000 vs. 1.19–8.36) followed similar trajectories, with a modest upward trend that became more pronounced in females after 2014. The lowest incidence observed in this age group was in Thailand, based on administrative claims data, where both sexes showed comparable trajectories. Incidence was highest in males (0.72–1.47 per 100,000), peaking in 2018 and 2020, while female incidence ranged from 0.33–0.83. Data for individuals aged <60 years were not available for the US Medicare population.

#### Individuals aged ≥60 years

Among individuals aged ≥60 years, pulmonary NTM incidence was consistently higher than in younger age groups across all regions. In Japan, incidence increased over time in both datasets. Among females, NDB-derived incidence increased from 112.70–123.80 per 100,000, while laboratory estimates ranged from 110.0–141.6, peaking in 2017. In males, laboratory-based incidence (101.9–127.6 per 100,000) was substantially higher than NDB estimates (30.4–33.2), highlighting divergence between data sources for this older age group. In the US Medicare population, incidence increased over time for both sexes. Incidence in females increased from 28.23–44.75 per 100,000, peaking in 2016, whereas incidence in males increased more gradually (18.0–22.51). In Queensland, Australia, incidence fluctuated over time for both sexes. Female incidence increased rapidly from 18.32–116.37 per 100,000, whereas male incidence increased more gradually (6.78–34.26). Conversely, in Thailand, incidence was higher in males (2.53–4.6 per 100,000) than females (1.09–3.28) in the older population, although rates remained substantially lower than in the high-income regions. Incidence increased steadily in males until 2020, with a marked decrease in 2022, while female incidence showed greater year-to-year variability.

### NTM species heterogeneity

The number of NTM species or species groups varied across regions, with 72 species detected in Queensland, 12 in Phnom Penh, 75 in Japan, and 10 in Thailand’s ODPC 5 region. Species identification practices differed, and some isolates lacked species-level identification (e.g. “NOT-TYPED”). In Queensland and Japan, the slow-growing MAC accounted for most cases, representing 53.24% and 77.77% of the five most frequently identified taxa, respectively. In Queensland, *M. intracellulare* predominated (40.17% of isolates), followed by *M. avium* (10.28%). In Japan, however, *M. avium* predominated (51.91%), followed by *M. intracellulare* (25.86%). *M. abscessus* was also consistently identified (8.48% of isolates in Queensland and 4.35% in Japan). In contrast, species distributions in Phnom Penh, Cambodia and Thailand’s ODPC 5 region differed. In Phnom Penh, rapid growers dominated (37.24% of the five most frequently identified taxa) with *M. abscessus* the most frequently identified species (21.94% of isolates), followed by the MFG (15.31%). In Thailand’s ODPC 5 region, *M. abscessus* was also the predominant species (18.51% of isolates), followed by *M. intracellulare* (12.93%). Across all four regions, MAC species, *M. abscessus*, and MFG species were consistently represented among the most frequently identified taxa, although their relative proportions varied.

## DISCUSSION

In this multi-region analysis, pulmonary NTM incidence increased across all regions, although incidence magnitude varied substantially by region and data source. Higher incidence was observed in the high-income regions of Queensland (Australia), Japan, and the US, whereas estimates were considerably lower in the middle-income regions of Phnom Penh (Cambodia) and Thailand. Upward trends were observed across both data sources, consistent with increasing detection of NTM pulmonary infections across diverse geographic settings. Because NTM pulmonary infections remain non-notifiable in most countries, the global burden is difficult to estimate. Nevertheless, epidemiologic studies consistently report increasing incidence of both NTM pulmonary infection and NTM-PD across multiple countries and data sources.^10,14^ Similar increases have been reported in high-income regions, including long-term surveillance studies in Queensland,^11,26^ nationwide analyses in Japan,^13,27^ and population-based studies in the US.^12,28^

Differences between administrative claims and laboratory data were apparent, particularly in Japan, where laboratory estimates consistently exceeded those derived from the NDB. This likely reflects differences in case definitions and ascertainment. Laboratory data capture microbiologically confirmed infection, whereas claims-based data capture clinically recognised disease.^13,14^ Laboratory systems may therefore include cases that do not meet clinical disease criteria.

Across high-income regions, NTM pulmonary infections showed strong age-dependent and sex-specific patterns, with increasing incidence in older adults and a shift toward female predominance. In Queensland, female predominance emerged after 2014 and was driven by higher incidence among older females. This pattern is consistent with prior reports describing a transition from cavitary disease in middle-aged males who smoked to nodular bronchiectasis-associated disease in elderly females.^29^ In Japan, similar age-dependent increases have been reported,^13^ differences between claims and laboratory data were observed in older males. This may partly reflect over-registration of elderly males in laboratory-based datasets, including cases where NTM was isolated during TB evaluation without follow-up or treatment. Sex-specific comorbidity profiles support this heterogeneity, with chronic kidney disease, osteoporosis, and Sjögren syndrome associated with NTM infection in females, and COPD and TB sequelae associated with infection in males.^30^

In contrast, demographic patterns differed in the middle-income regions. In Thailand, incidence was higher among individuals aged ≥60 years, with persistent male predominance, likely reflecting a greater burden of prior TB and structural lung disease. Similar male predominance in NTM pulmonary infection has been reported in other high TB-burden regions.^31^ Prior TB infection is an important risk factor for subsequent NTM-PD due to residual lung damage and bronchiectasis.^32^ In Phnom Penh, incidence remained low and no consistent sex predominance was observed. Large year-to-year variability likely reflects differences in case detection, diagnostic practices and TB screening activity rather than true epidemiologic trends. Under-ascertainment may be compounded by increasing reliance on molecular TB diagnosis, which do not identify NTM and reduce use of mycobacterial culture required for NTM detection. In a previous Cambodian study, 60.6% of TB cases were clinically diagnosed without mycobacterial culture,^33^ limiting NTM detection and identification of TB–NTM co-infections.

NTM species distributions also differed between regions. Species diversity was greater in Queensland and Japan than in Phnom Penh and Thailand, likely reflecting broader diagnostic coverage. In Queensland and Japan, MAC predominated, particularly *M. intracellulare* in Queensland and *M. avium* in Japan. MAC species account for the majority of pulmonary NTM infections in these high-income regions.^11,13,26,27,34^ In contrast, rapidly growing mycobacteria comprised a larger proportion of isolates in Phnom Penh and Thailand’s ODPC 5 region, where *M. abscessus* was most frequently identified. These findings are consistent with studies from tropical and TB-endemic regions in Asia reporting high case numbers of rapidly growing mycobacteria, particularly *M. abscessus*.^35–37^ Regional differences may reflect environmental exposures, including climate-related factors, given higher rates of *M. abscessus* in tropical regions.^11,26,37–39^ They may also reflect differences in host risk factors, diagnostic practices, laboratory capacity, and species-level identification.

Several limitations should be considered. Case definitions differed across datasets, including incident case criteria, laboratory reporting practices (e.g., calendar-year vs. ≥12-month definitions), and administrative coding, limiting direct comparisons of absolute incidence. Observation periods also differed, with Queensland contributing more than 24 years of surveillance data, which may affect trend estimates relative to regions with shorter follow-up. Changes in laboratory identification methods, including introduction of MALDI-TOF and updates to PCR-based genotyping, may have affected species distribution. In Cambodia, incidence estimates were derived from a single testing centre in Phnom Penh but standardised to the national population, likely underestimating true incidence. In TB-endemic regions, diagnostic pathways may prioritise molecular TB testing, reducing culture use required for NTM identification. This TB prioritisation may also contribute to the male predominance observed in Thailand, as screening disproportionately captures men with respiratory symptoms and comorbidities. In the US, estimates were restricted to Medicare beneficiaries aged ≥65 years, limiting comparisons with younger populations. In Japan, claims-based analyses defined incident cases by the absence of prior claims within the available lookback period. As this period lengthened over time, prior disease was more likely to be identified, increasing classification as prevalent rather than incident. This may have resulted in progressive underestimation of incidence in later years. Further, incidence estimates were not directly comparable with other regions, as they were derived using an extrapolation method based on the ratio of NTM cases to pulmonary TB notifications, rather than population-based denominators. As this method was developed in the 1970s, changes in TB epidemiology and population demographics mean these estimates should be interpreted with caution.^40–42^

## CONCLUSION

Pulmonary NTM incidence increased across all regions, although magnitude varied by health care system and diagnostic practices. Higher incidence in high-income regions likely reflects more consistent diagnostic practices, whereas lower estimates in middle-income regions likely reflect under-ascertainment related to diagnostic access, healthcare infrastructure, and TB prioritisation. In TB-endemic settings, prioritisation of TB diagnosis and control further contribute to under-recognition of NTM pulmonary infection. Geographic differences in species distribution were evident, with MAC predominating in Japan and Queensland, while *M. abscessus* predominated in tropical regions. These findings illustrate the challenges of comparing NTM epidemiology across regions and highlight the importance of strengthening diagnostic capacity and surveillance to better define the global burden of NTM pulmonary infection.

## Ethics approval Australia

This study was approved by The Prince Charles Hospital Human Research Ethics Committee (HREC/15/QPCH/65). Approval to access NTM notification data was granted by Queensland Health (27466) under the Public Health Act 2005., **Cambodia:** This study was approved by the National Ethical Committee for Health Research (NECHR) on September 17, 2024 (Ref. No. 297 NECHR)., **Japan:** The study protocol was approved by the Institutional Review Board of Fukujuji Hospital (Approval No. 23040)., **Thailand:** This study was approved by the Ethical Review Committee for Research in Human Subjects, Ministry of Public Health, Thailand (Document No. 1/2025; Protocol Ref. No. 20/2567), and the Ethics Committee for Research in Human Subjects, Department of Disease Control, Ministry of Public Health, Thailand (FWA 00013622). The study used anonymized Electronic Medical Records (43 Files Standard Data Set) obtained from the Health Data Center and laboratory data from the Office of Disease Prevention and Control Region 5., **United States:** All methods were carried out in accordance with relevant guidelines and regulations of the National Institutes of Health’s Office of Human Subjects Research Protection. Analysis of the data was deemed “Not Human Subjects Research”, under the 2018 Health and Human Services Common Rule because of the use of deidentified data and, accordingly, is exempt from Institutional Review Board review.

## Author contributions

Conceptualisation: ARP, MMA, RAM, DRP, RMT

Funding acquisition: SC, KM, SM, DRP, RMT

Supervision: SC, KM, SM, DRP, MMA, RMT

Project administration: RAM, LK, EM, SU, PW, FG

Data curation: EMT, EM, SU, CY, KK, SW, PT, NT, SP, PW, RAM, FG, MMA, ARP

Methodology: DRP, RMT, MMA, ARP

Formal analysis: MMA, ARP

Visualisation: MMA, ARP

Writing – original draft: MMA, ARP

Writing – editing: SC, SU, CY, PW, SM, LK, EM, KM, RAM, DRP, RMT, MMA, ARP

All authors reviewed the manuscript and approved the final version for submission.

## Declaration of Generative AI and AI-assisted technologies in the writing process

No AI tools were used in the conduct of the study, analysis of the data, or preparation of the manuscript.

## Data availability

The data used in this study were obtained from multiple country- and region-specific administrative and laboratory databases and are subject to institutional, governmental, and data-use restrictions and therefore cannot be shared publicly, including in de-identified form. Requests for access to aggregated data may be requested from the relevant data custodians, subject to local ethical approvals and data governance requirements.

## Abbreviations

AFB: acid-fast bacilli
ATS: American Thoracic Society
CI: confidence interval
CMS: Centers for Medicare and Medicaid Services
HDC: Health Data Center
ICD: International Classification of Diseases
IDSA: Infectious Diseases Society of America
IRR: incidence rate ratio
LMICs: low- and middle-income countries
MAC: *Mycobacterium avium* complex
MALDI-TOF: Matrix-Assisted Laser Desorption/Ionization Time-of-Flight
MFG: *Mycobacterium fortuitum* Group
NDB: National Health Insurance Claim Database
NoCS: Notifiable Conditions
NTM: Nontuberculous mycobacteria
ODPC 5: Office of Disease Prevention and Control Region 5
PD: pulmonary disease
PNH: Phnom Penh
QLD: Queensland
TB: Tuberculosis
US: United States

## Acknowledgements

**Australia:** We would like to thank Bridget O’Connor for her assistance in supplying the Queensland Health Notifiable Conditions Database.

**Thailand:** We thank Sarrote Boonkerd for consultation on the Electronic Medical Records system (43 Files Standard Data Set) at the Health Data Center, Ministry of Public Health; Charuttaporn Jitpeera for epidemiological advice; and Natnicha Ritticot, Sumet Amonyingcharoen, and Lapasrada Pattarapreeyakul for their valuable insights on infectious diseases.

